# Methylprednisolone as Adjunctive Therapy for Large Vessel Occlusion (MARVEL): a Randomized Double-blind, Placebo-controlled Trial in Patients undergoing Endovascular therapy: Study Rationale and Design

**DOI:** 10.1101/2023.08.02.23293575

**Authors:** Qing-Wu Yang, Changwei Guo, Chengsong Yue, Jie Yang, Linyu Li, Zhouzhou Peng, Jinrong Hu, Jiandi Huang, Jiaxing Song, Jiacheng Huang, Weilin Kong, Nizhen Yu, Dahong Yang, Xiang Liu, Duolao Wang, Raul G. Nogueira, Fengli Li, Thanh N. Nguyen, Wenjie Zi

## Abstract

**Background:** Steroids have pleiotropic neuroprotective actions including the regulation of inflammation and apoptosis which may influence the effects of ischemia on neurons, glial cells, and blood vessels. The effect of low-dose methylprednisolone in acute ischemic stroke (AIS) patients in the endovascular treatment (EVT) era remains unknown.

**Objective:** This trial investigates the efficacy and safety of low-dose methylprednisolone (2 mg/kg intravenous for 3 days) as adjunctive therapy for AIS patients undergoing mechanical thrombectomy (MT) within 24 hours from symptom onset.

**Methods and design:** Methylprednisolone as Adjunctive Therapy for Acute Large Vessel Occlusion (MARVEL) Trial is an investigator-initiated, prospective, randomized, double-blind, placebo-controlled multicenter clinical trial. Up to 1672 eligible patients with anterior circulation large vessel occlusion (LVO) stroke presenting within 24 hours from symptom onset are planned to be consecutively randomized to receive methylprednisolone or placebo in a 1:1 ratio across 82 stroke centers in China.

**Outcomes:** The primary outcome is the ordinal shift in the modified Rankin scale (mRS) score at 90 days. Secondary outcomes include 90-day functional independence (mRS 0-2). The primary safety endpoints include mortality at 90 days and symptomatic intracerebral hemorrhage within 48 hours of MT.

**Conclusion:** The MARVEL trial will provide evidence of the efficacy and safety of low-dose methylprednisolone as adjunctive therapy to anterior circulation LVO stroke patients undergoing endovascular treatment.

**Trial registry number:** ChiCTR2100051729 (www.chictr.org.cn).

## Introduction

The American Heart Association (AHA) guidelines recommend endovascular treatment (EVT) for select LVO patients with class IA evidence. The recanalization rate has improved to 80%-94% with newer endovascular techniques.^1, 2^ As high recanalization rates have been achieved, research priorities pivot toward neuroprotection to improve patient outcomes. Theoretically, neuroprotection can be achieved either by extending the longevity of the penumbral territory that could be salvaged, limiting hemorrhagic and edema risks with reperfusion, or attenuating the potential risks of reperfusion injury and related inflammatory cascade post-reperfusion. Intercepting any of these pathways may help to decrease the risk of “futile recanalization”, a term that describes patients who achieve satisfactory reperfusion but fail to achieve functional improvement.^3–9^ Another term that has recently emerged in lieu of “futile recanalization” is “reperfusion without functional independence” because many of these patients can still have quality of life despite being in a state of dependency. ^10^

Growing evidence indicates that the inflammatory response following stroke are present throughout the brain and can worsen secondary brain injury.^11^ Cerebral edema after ischemic stroke can be classified as cytotoxic or vasogenic edema. While most of the edema after ischemic infarction is vasogenic, cytotoxic edema can also occur due to cell membrane dysfunction. Several early phase I/II trials focused on adjunctive immune-targeted therapies to EVT have yielded promising results of reducing disability and mortality compared to control patients.^4, 12, 13^

While corticosteroids are widely used anti-inflammatory and immunosuppressive agents, their effect on stroke outcomes remains debated. Preclinical studies have shown that corticosteroids can reduce infarct area in animal models^14, 15^ while clinical stroke studies have shown neutral results.^16^ However, there was a call to reignite the study of corticosteroids in stroke, especially in the reperfusion era. ^17, 18^ Several lessons are learned from prior studies. First, previous neutral studies used high doses and a long duration of corticosteroid which may have led to more side effects. Second, the sample size of these trials was small and most included patients with “presumed ischemic stroke” were treated in an era where reperfusion was not effective. Davis and Donnan suggested that steroid therapy for stroke be discarded prematurely. Third, the last trial was published in 2001 by Ogun et. al.^19^ To date, the evidence for the use of corticosteroids in stroke is limited. High quality evidence is warranted to provide a definitive answer to the benefit or risk of adjunctive therapy of corticosteroids in patients with stroke. Fourth, endovascular treatment has transformed the treatment of ischemic stroke patients with large vessel occlusion. While 80% to 90% of patients achieve reperfusion, approximately half of patients are disabled and the mortality is estimated 15%.^20^ Improving functional outcome, reducing disability and mortality for patients who have achieved reperfusion is an important frontier in stroke research.^21^ Thus, adjuvant treatments are needed to complement recanalization therapies.^22^ ^17^Considering the reasons above, studies with adequate sample size are warranted to give a high-level evidence to prove or disprove the use of corticosteroids in the new reperfusion era.

We initiated the MARVEL trial to give a high level of evidence of whether use of low-dose methylprednisolone, an intermediate-acting corticosteroid with a low side effect profile and a strong ability to penetrate the brain-brain barrier, can benefit or not benefit stroke patients in the era of EVT in a large, multicenter, double-blind, randomized clinical trial.^23^

## Methods

### Design

The Methylprednisolone as Adjunctive Therapy for Acute Large Vessel Occlusion (MARVEL) trial is a multicenter, prospective, randomized, placebo-controlled, double-blind clinical trial. This trial was registered at www.chictr.org.cn (ChiCTR2100051729). The trial was designed in compliance with the Declaration of Helsinki. The protocol was approved by the ethics committee of Chongqing Xinqiao Hospital, Army Medical University, and all participating centers. This study included 82 stroke centers in China. The key criterion for a qualifying participating center is the performance of at least 30 mechanical thrombectomy procedures with the use of stent-retriever or contact aspiration devices annually, and all participating neuro-interventionalists should have more than 3 years experience of neuro-intervention and performed at least 30 EVT. The trial flowchart is depicted in Figure 1.

**Figure 1.**
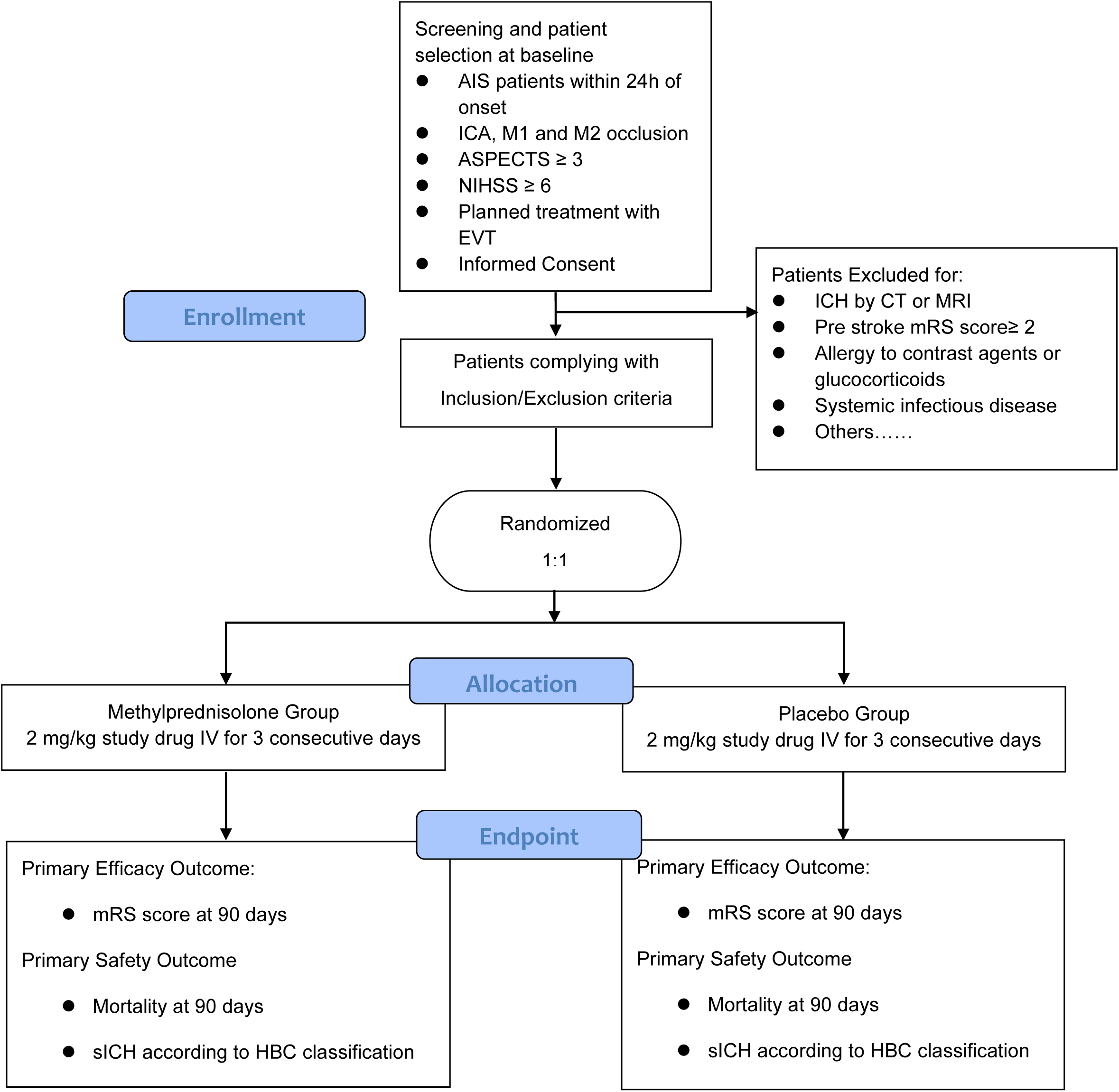
Study flowchart. Abbreviations: AIS, acute ischemic stroke; ICA, intracranial carotid artery; M1, first segment of the middle cerebral artery; M2, second segment of the middle cerebral artery; ASPECTS, Alberta Stroke Program Early CT score; NIHSS: National Institute of Health Stroke Scale; EVT, endovascular treatment; ICH, intracranial cerebral hemorrhage; CT, computed tomography; MRI, magnetic resonance imaging; mRS, modified Rankin Scale; sICH, symptomatic intracranial hemorrhage; HBC, Heidelberg Bleeding Classification

Inclusion criteria include:

1. Age ≥ 18 years
2. Presenting with AIS with symptoms within 24 hours from time last known well
3. Baseline National Institutes of Health Stroke Scale (NIHSS) ≥ 6
4. Anterior circulation ischemic stroke was determined according to clinical symptoms and imaging examination
5. Baseline Alberta Stroke Program Early CT Score (ASPECTS) ≥ 3
6. Occlusion of the intracranial internal carotid artery, the M1- or M2-segment of the middle cerebral artery confirmed by CT, MR angiography, or digital subtraction angiography
7. Planned treatment with EVT
8. Informed consent obtained from patients or their legal representatives.

Exclusion criteria include:

1. CT or MRI evidence of hemorrhage
2. mRS score ≥ 2 before stroke onset
3. Pregnant or lactating women
4. Allergic to contrast agents or glucocorticoids
5. Participating in other clinical trials
6. Dual antiplatelet treatment within 1-week prior to onset
7. Systolic blood pressure > 185 mmHg or diastolic pressure > 110 mmHg, refractory to antihypertensive drugs
8. Genetic or acquired bleeding diathesis, lack of anticoagulant factors, or oral anticoagulants and INR > 1.7
9. Blood glucose < 2.8 mmol/L (50 mg/dl) or > 22.2 mmol/L (400 mg/dl), platelets < 90 × 10^9/L
10. Arterial tortuosity precluding access with thrombectomy device
11. Bleeding history (gastrointestinal and urinary tract bleeding) in prior 1 month
12. Chronic hemodialysis and severe renal insufficiency (glomerular filtration rate < 30 ml/min or serum creatinine > 220 umol/L [2.5 mg/ dL])
13. Life expectancy < 6 months due to comorbidities
14. Follow-up is not expected to be completed
15. Intracranial aneurysm or arteriovenous malformation
16. Brain tumor with mass effect on imaging
17. Systemic infectious disease

### Randomization

After confirming patient eligibility, randomization will be conducted through a web-based application (www.jinlingshu.com) stratified by participating centers. Eligible patients will be randomly assigned to either the methylprednisolone or control group in a 1:1 ratio. Patients enrolled in the trial will receive masked medications corresponding to their assigned random serial number based on their enrollment time. Both trial personnel and patients will remain blinded to treatment assignment throughout the study.

### Contents of study drug kit

Each kit contains 12 bottles of drugs, and each bottle contains 40mg of lyophilization of methylprednisolone sodium-succinate or placebo. The drug is manufactured and provided by Chongqing Lummy Pharmaceutical Co., Ltd. The medication kit and bottles are visually identical (including labeling, dosage form, size, and color), except for the identification number.

### Treatment

After randomization, the study drug will be dissolved in saline with a dose of 2mg/kg intravenous (IV) per day and administered immediately upon patient enrollment. The maximum dose is 160mg IV. The study drug will be given for three consecutive days and then ceased. Gastrointestinal prophylaxis and hyperglycemia treatment will be administered per standard local protocols.

### Efficacy endpoints

The primary endpoint is the distribution of the modified Rankin Scale (mRS, shift analysis) at 90 days after randomization.

The secondary endpoints include:

1. proportion of mRS score 0 to 4 at 90 days,
2. proportion of mRS score 0 to 3 at 90 days,
3. proportion of mRS score 0 to 2 at 90 days,
4. proportion of mRS score 0 to 1 or return to pre-morbid mRS score at 90 days (for patients with mRS >1)
5. European Quality Five-Dimension scale score at 90 days.

### Safety endpoints

The safety endpoints include mortality at 90 days, symptomatic intracranial hemorrhage (SICH) rate within 48 hours according to the Heidelberg Bleeding Classification,^24^ the proportion of patients with any intracranial hemorrhage within 48 hours, or hemicraniectomy. The incidence of serious adverse events and corticosteroid-related adverse events (hyperglycemia, infection, and gastrointestinal hemorrhage) will also be recorded.

### Data and safety monitoring board (DSMB)

An independent DSMB will be organized with 3 experts (including a neurologist, a neuro-interventionist, and a biostatistician). Members of the DSMB will not participate in the trial or be affiliated with the study sponsors. The DSMB will meet annually and monitor trial progress. In addition, the DSMB will review the incidence of serious adverse events to ensure patient safety.

### Sample size estimates

We assume favorable functional outcome (mRS 0-2) rates of 50% and 43% in the methylprednisolone and placebo groups, respectively. ^25, 26^ The steering committee estimated a 7% difference in improved outcomes with steroids. To demonstrate a 7% absolute difference with a type-1 error alpha of 0.05 (two-tailed) with a power of 80% (beta 20%), a sample size of 1588 patients would be needed (794 patients per treatment group). To take into account a 5% attrition rate, a total of 1672 patients is required (836 per treatment group).

### Statistical analysis

The primary effect variable will be the common odds ratio (cOR) and will be estimated with a generalized linear model (GLM) if the odds proportion assumption is satisfied. Otherwise, an assumption-free method will be used to calculate the generalized odds ratio.^27^ The secondary outcomes and safety outcomes will be analyzed using GLMs. The treatment effect will be estimated using GLMs adjusted for the following prognostic variables a priori: age, baseline NIHSS score, pre-stroke mRS, baseline ASPECTS score, use of intravenous thrombolysis, time from onset to randomization, and occlusion location. Primary data analyses will be based on the intention-to-treat principle. The per-protocol analyses will also be performed as supplemental analysis. All statistical analyses will be performed using SAS 9.4 and R 4.3.0. The trial results will be reported following the CONSORT guidelines for reporting randomized trials.

### Discussion and summary

The MARVEL trial is planned to involve 1672 patients which is nearly four-fold higher than the Cochran review of 466 patients.

Evidence-based treatments to salvage the penumbral tissue invariably involve restoring blood flow as early as possible.^28^ Revascularization strategies including intravenous thrombolysis (IVT) and EVT are recommended in select patients by multiple neuroscience organizations.^1, 29, 30^ However, while the early recanalization rate is estimated to be achieved in more than 80% in EVT patients, about half of patients do not achieve functional independence.^10, 20, 31^ Several studies indicated that modulating the immune system could be a target to close this gap in patient outcomes.^12, 32, 33^

Inflammatory mechanisms after stroke are now increasingly considered prime targets for stroke therapy since immune signals and their mediators can have both detrimental and beneficial effects at different stages of the disease process.^11, 32^ Several proof-of-concept trials have demonstrated that immune modulators were effective in AIS patients undergoing EVT.^3, 4, 12, 34^ Adjunctive therapies are now being increasingly considered to complement endovascular treatment. Shi’s Team suggested that fingolimod may be a potential treatment for ischemic stroke patients. The Phase Ib/IIa APRIL trial investigated a DNA aptamer Aptoll which is an antagonist of toll-like receptor 4 (TLR4), as an adjunctive therapy to EVT. The results were promising, indicating reduction in disability and mortality at 90 days.^13^ However, the high cost and limited availability of this treatment restrict its widespread application. Corticosteroids, being widely used and easily accessible anti-inflammatory and immunosuppressive agents, have demonstrated their ability to reduce the area of infarction in animal models. It has been suggested that corticosteroids can restore the overall structure and improve the tightness of the BBB. (16)^15, 35^ However, the use of corticosteroids in clinical practice remains a subject of debate as evidence for its benefit in AIS patients remains sparse.^36^

A Cochrane Review on steroids for AIS evaluated 8 trials with 466 patients.^16^ There was bias in the selection of patients with “presumed ischemic stroke”. Since 2001, there has been no clinical trial dedicated to the use of corticosteroids in the management of patients with AIS in the reperfusion era.

The AHA guidelines do not recommend conventional or large doses of steroids for stroke patients due to a lack of evidence of efficacy and the potential to increase the risk of infectious complications. However, previously reported negative results did not deter physicians from the United States or China from administering corticosteroids to stroke patients, wherein approximately 20% of physicians surveyed reported routine steroid use.^16, 37, 38^ There is a need for high-quality evidence with sufficient sample size and lower doses of corticosteroids to determine whether or not to administer steroids to stroke patients.

The RESCUE-JAPAN-LIMT, ANGEL-ASPECTS, and SELECT2 trials have expanded the use of EVT to treat patients with large infarct cores.(Ref) It has been suggested that corticosteroids may be beneficial for high-risk patients with large infarcts and significant vasogenic edema, and may not be effective or could even be harmful for those with smaller infarct and less vasogenic edema as suggested by Mulley. This study includes broad imaging eligibility criteria such as a baseline ASPECTS score ≥ 3, which is consistent with recent evidence to treat patients with small and large infarct core. Additionally, this study aims to investigate the effect of corticosteroids on patients experiencing high severity of stroke disease events associated with high morbidity and mortality.

Hence, we initiated this randomized trial to learn whether a widely available clinical immune modulation agent, methylprednisolone, confers benefit as adjunctive therapy to AIS patients undergoing EVT. The MARVEL randomized trial enrolled the first patient on 09 February 2020. When completed, this trial will provide pivotal data allowing an assessment of the efficacy and safety of early adjunctive low-dose methylprednisolone with EVT for patients with AIS.

## Data Availability

The data that support the findings ofthis study are available from thecorresponding author upon reasonable request.

---

**Table 1.**
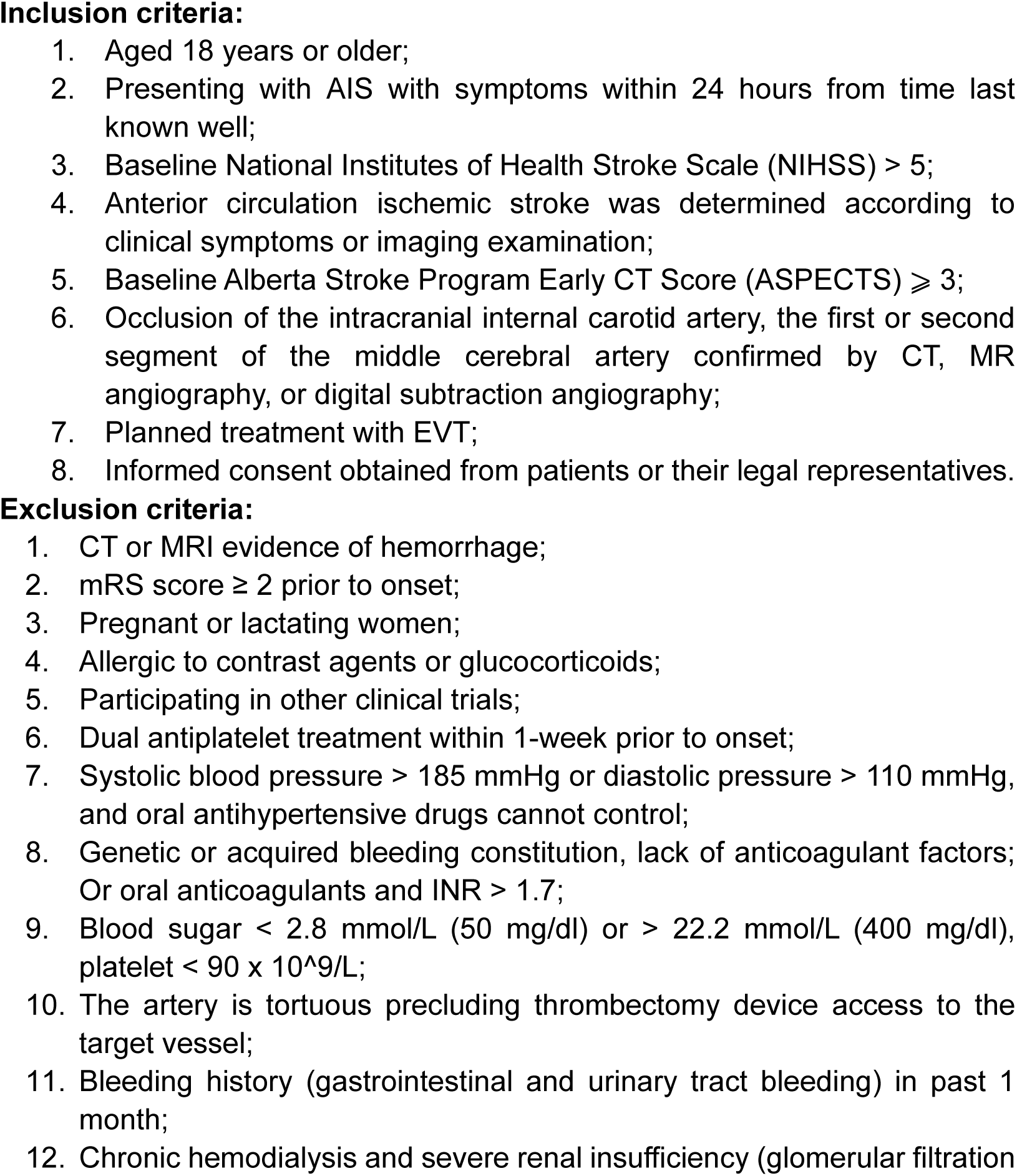
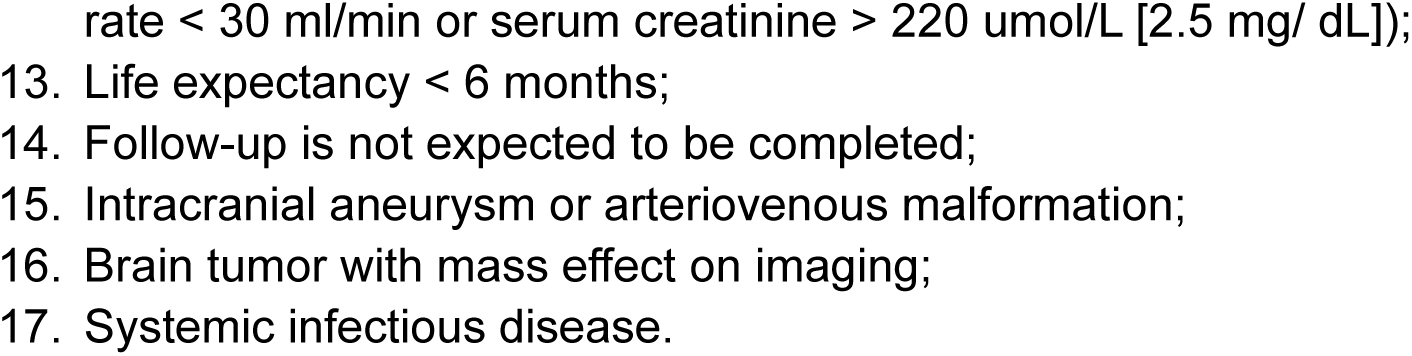
MARVEL Trial Inclusion and Exclusion Criteri.

